# Using Genetics, Genomics, and Transcriptomics to Identify Therapeutic Targets in Juvenile Idiopathic Arthritis

**DOI:** 10.1101/2024.11.01.24316521

**Authors:** Evan Tarbell, James N. Jarvis

## Abstract

Despite progress in improving outcomes for oligoarticular and polyarticular juvenile idiopathic arthritis (JIA), the field still faces considerable challenges. More than half of adults who had JIA continue to have active disease and have developed functional limitations. Medication side-effects are common and intrusive. Thus, the field continues to search for therapeutic agents that target specific aspects of disease pathobiology and will be accompanied by fewer and less intrusive side effects. We identified 28 candidate target genes that were associated with JIA according to Open Targets Genetics and were also differentially expressed in the CD4+ T cells of children with active JIA patients (when compared to healthy controls). Of the 28 candidates, the strongest new target to emerge was homeodomain interacting kinase (HIPK1), which suppresses T cell activation and is within the *PTPN22* locus tagged by rs6679677. This locus includes an enhancer element that contacts the HIPK1 promoter, and HIPK1 shows decreased expression in JIA CD4+ T cells when compared to controls. Gene Ontology terms associated with HIPK1 were overrepresented among the differentially expressed genes between JIA and controls and PML, a known co-regulator of HIPK1, showed a similar suppressed gene expression profile. Two downstream transcription factors of HIPK1, TP53 and GATA4, showed enriched binding patterns near the promoters of JIA up-regulated genes. Taken together, these data suggest a pathogenic role for HIPK1 in JIA and make it a prime candidate for therapeutic modulation.

## Introduction

Juvenile idiopathic arthritis (JIA) is a chronic condition characterized by chronic inflammation of the synovium; it is one of the most common chronic conditions affecting children in the USA^1,2^. Considerable progress that has been made in treating children with JIA over the past 20 years, particularly with the introduction of biological agents^3^. However, the field still faces important challenges. Despite progress in improving functional outcomes, these outcomes have come at a cost, as most children experience significant and intrusive medication side-effects from the initiation of treatment^4^. Furthermore, we face additional serious challenges: (1) we have had limited success in getting children off medication^5^, meaning that children with JIA have prolonged exposure to potent immunosuppressive medications; and (2) half of all adults diagnosed with JIA continue to have active disease and significant functional limitations, which results in exposure to these medications, and their toxicities/side-effects, that continue into adulthood^6^.

In the past 5 years, investigators have assessed the potential of using genetic, genomic, and transcriptomic data to guide the development of new therapies or to repurpose known agents^7, 8^. The underlying premise of these investigations is that these data sets, used together, are likely to identify features of pathobiology that are not discernable from any single measurement/assessment. Furthermore, by elucidating genetic risk and the immunologic pathways influenced by risk-inducing variants, there is reason to believe that personalized approaches to therapy can be provided to individual patients.

In this paper, we demonstrate the feasibility of using the known JIA genetic risk loci, 3D chromatin data, and RNA sequencing to identify candidate therapeutic targets. We identified twenty-eight candidate targets, one of which, HIPK1, is novel and represents a potentially druggable pathway.

## Materials and Methods

*<u>Patients and controls</u> -* Patient data (RNAseq, ChIPseq, HiChIP, and ATAC seq) used for these studies were previously reported in an earlier paper from our group^9^. We repeat some detail on the patient characteristics here. Patients fit criteria for polyarticular-onset, rheumatoid factor-negative JIA as established by the International League Against Rheumatism (ILAR)^46^. We used the Wallace criteria ^32–34^ to characterize children as having active disease on therapy (ADT) or clinical remission on medication (CRM). Patients with JIA were all being treated with combinations of methotrexate and the TNF inhibitor, etanercept. Samples were obtained from children classified as ADT and were obtained 6 weeks from the onset of therapy. Samples obtained from children with CRM were obtained when CRM status was confirmed. This was typically 12-15 months after the initial diagnosis.

Healthy control (HC) children were recruited from the Hodge General Pediatrics Clinic of the University at Buffalo Jacobs School of Medicine and Biomedical Sciences. Exclusion criteria fever ≥38°C within the previous 48 hr., presence of another autoimmune disease (e.g., type 1 diabetes), treatment with systemic glucocorticoids or antibiotics.

### Laboratory Data

Data from RNAseq and ChIPseq/HiChIP for CTCF in purified CD4+ T cells is available on the Gene Expression Omnibus under accession number GSE164215, uploaded in January, 2021 and updated in April, 2021. This data were used for the analyses described below. The STRING-DB database^19^ for protein-protein interactions was utilized to identify the interacting partners to HIPK1. Each member of the interaction network was used as a search term for FactorBook^20^, a repository of publicly available transcription factor binding data. Two of the interacting proteins were identified as transcription factors, TP53 and GATA4. FactorBook listed two TP53 ChIP-Seq datasets (accessions ENCFF849LSO in HepG2 cells and ENCFF699UTZ in A549 cell line) and one for GATA4 (accession ENCFF154VTV in HepG2 cells), which were downloaded.

### Data Analysis

#### Processing RNA-Seq Data

All RNA-Seq data analysis was performed on the Galaxy bioinformatics online platform^21^. Raw RNA-Seq FASTQ files were downloaded from the SRA archive^22^ using the fastq-dump tool^23^. The paired-end FASTQ files were aligned to the hg38 genome using RNA-STAR^24^, using the GENCODE^25^ Version 44 GTF file as the gene model for splice junctions and with the “GeneCounts” option set to create a table of read counts per gene. All other parameter settings were left to their Galaxy defaults. These count table files were used as the input to the edgeR differential expression analysis tool^26^, which was run without low count genes, defined as having a cpm value below 1.0 in less than 10 samples, and with all other settings at their default values. Differentially expressed genes were used for GO term analysis using Gorilla^27^. GOrilla was run using the two gene set option, with the up-regulated or down-regulated genes as the input and the total list of expressed genes as the background set.

*Determining Regulatory Potential of Transcription Factors for Differentially Expressed Genes*

#### BETA^28^ was used to calculate the regulatory potential of the HIPK1-associated transcription factors for the differentially expressed genes in JIA T-cells

The regulatory potential, which is a gene’s likelihood of being regulated by a factor, is estimated for each gene. The regulatory potential is calculated as:

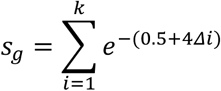

All binding sites (k) near the transcription start site of the gene (g) within a user specified range (100 kb as default) are considered. Δ is the exact distance between a binding site and the TSS proportional to 100 kb (Δ = 0.1 means the exact distance = 10 kb). BETA then generates a cumulative distribution function of the gene groups and uses a one-tailed Kolmogorov-Smirnov test to determine whether the Up-regulated and Down-regulated groups differ significantly from the Non-differential group.

## Results

In order to identify the pathogenic genes that are controlled by non-coding JIA-associated single nucleotide polymorphisms (SNPs) and then to determine which of those represented viable putative drug targets, the following approach was employed. First, the Open Targets Genetics (OTG) platform^29^ was used to link potential pathogenic genes to JIA-associated genetic variants. Those genes whose link to JIA was predicted by OTG were then cross referenced with genes that were determined to be differentially expressed in CD4+ T-cells between patients with active JIA and healthy controls, originally published by Tarbell et al.^9^ A total of 29 genes met this criteria (**Table 1**). With the focus on novel drug targets, genes were then excluded if there were known clinical-stage drugs that targeted them. Four of the 29 genes initially identified met this criteria: *ALDH2*, which has been targeted for alcohol disorders and has been tested for some cancers (**Supp Table 1**); *CHRNB2*, a target of nicotine; *ITGAL* (CD11a), was targeted for psoriasis, arthritis and diabetes (**Supp Table 1**) and; *ITGAM* (CD11b), frequently, although unsuccessfully, targeted for cancers^30^. The remaining 25 identified genes were then assessed for drug tractability, using the Open Targets Platform, which includes tractability data that identifies key details for small molecules, antibodies and other modalities. For small molecules, assessments include: whether the target has been co-crystallized with a small molecule, whether there is a high-quality ligand, whether there is a high-quality binding pocket, whether there is a med-quality pocket or if the target is part of a druggable family, as per Finan et al’s Druggable Genome pipeline^31^. For antibodies, the assessments include: whether there is high confidence that the subcellular location of the target is either plasma membrane, extracellular region/matrix, or secretion, that there is medium confidence that the subcellular location of the target is either plasma membrane, extracellular region/matrix, or secretion, that the target has a predicted signal peptide or trans-membrane regions, and not destined to organelles, or that there is high confidence that the target is located in the plasma membrane. Overall, 6 of the 25 candidate genes/gene products met at least one of these criteria for either small molecule or antibody tractability (**Table 2**). Of these top 6 candidates, the most promising one was Homeodomain Interacting Protein Kinase 1, or HIPK1, a putative effector gene of the JIA-associated SNP, rs6679677. The Open Targets Genetics platform was used to identify 10 putative effector genes of the rs6679677 SNP (**Table 3**). The highest scoring target gene from OTG was *PTPN22*, followed by the nearest gene in linear genomic space, *PHTF1*, and with HIPK1 rounding out the top 5. Of note is that *HIPK1*, although it had the fifth overall score assessed by OTG, had the third highest rank in eQTL colocalization scores and chromatin interaction scores, components of the OTG model that use experimental data. To further prioritize the putative targets from OTG, publicly available transcriptomic and genomic data in the CD4+ T cells of patients with JIA and age matched healthy controls, originally published by Tarbell et al.^9^, was downloaded from the SRA data repository and processed (See Methods). Transcriptomic data, in the form of RNA-seq, was pooled and averaged across the subjects in each disease group, as was genomic data in the form of ATAC-seq, CTCF ChIP-seq and HiChIP, and were graphically displayed in a genome browser, centered on the rs6679677 SNP (**Figure 1**). As shown in the genome browser screenshot, there are several regulatory regions in the vicinity of the SNP, denoted by spikes in the ATAC-seq signals. Those regulatory elements near the rs6679677 SNP are in close physical proximity to numerous distal regulatory elements, as shown with the HiChIP loops. Finally, many genes in the vicinity to the rs6679677 SNP, and linked to it via chromatin loops, show robust expression as shown in RNA-seq signal tracks.

**Figure 1.**
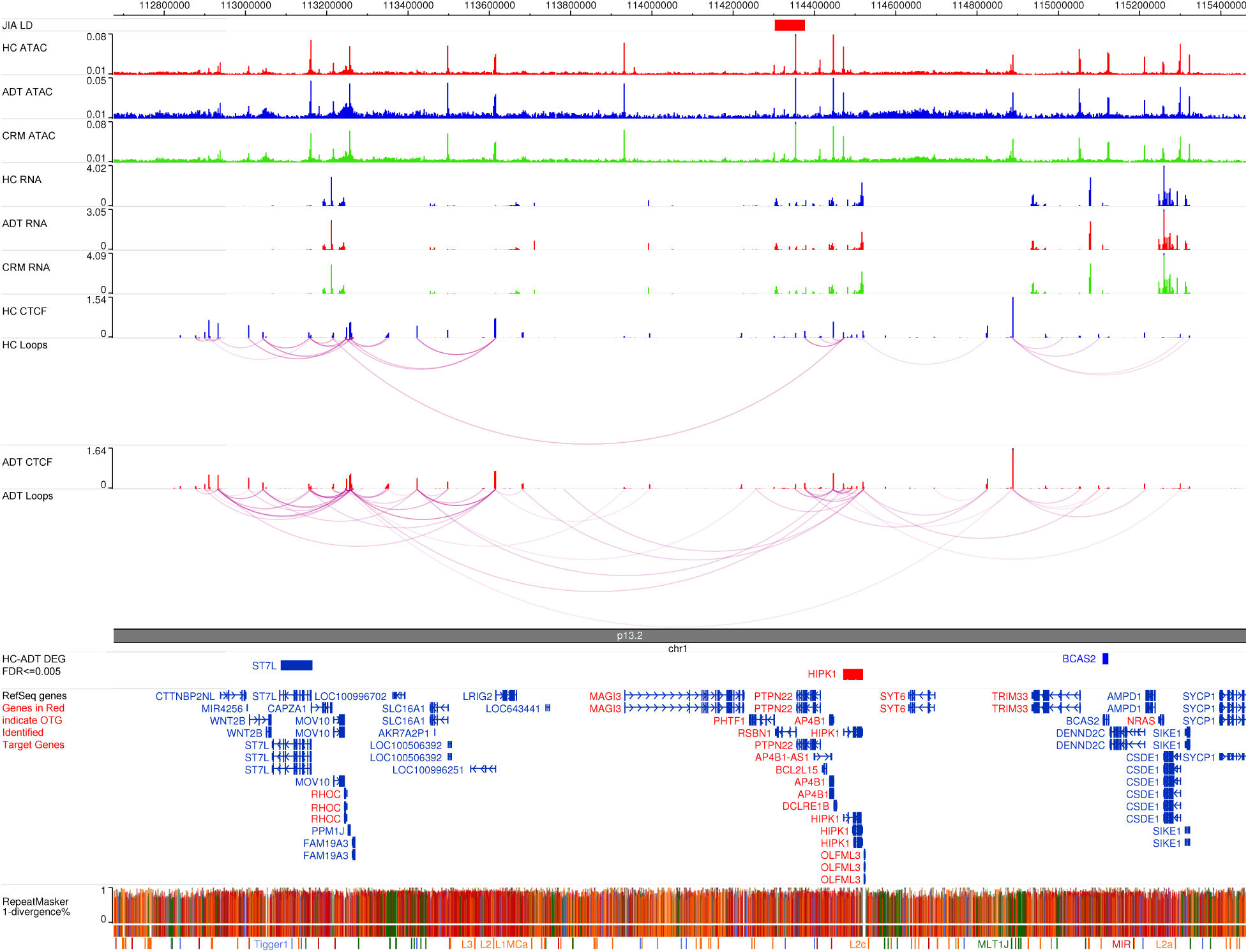
Genome Browser Snapshot of rs6679677 JIA associated SNP. Genome Browser snapshot shows data generated in T-cells of JIA patients (Active disease under treatment–ADT and clinical remission on medication–CRM) and matched healthy controls (HC). Pooled ATAC-seq, RNA-seq, CTCF ChIP-seq and CTCF HiChIP data is shown, as generated in Tarbell et al., 2021. Genes in red indicate genes identified by Open Targets Genetics as target genes of rs6679677 SNP.

**Table 1.**
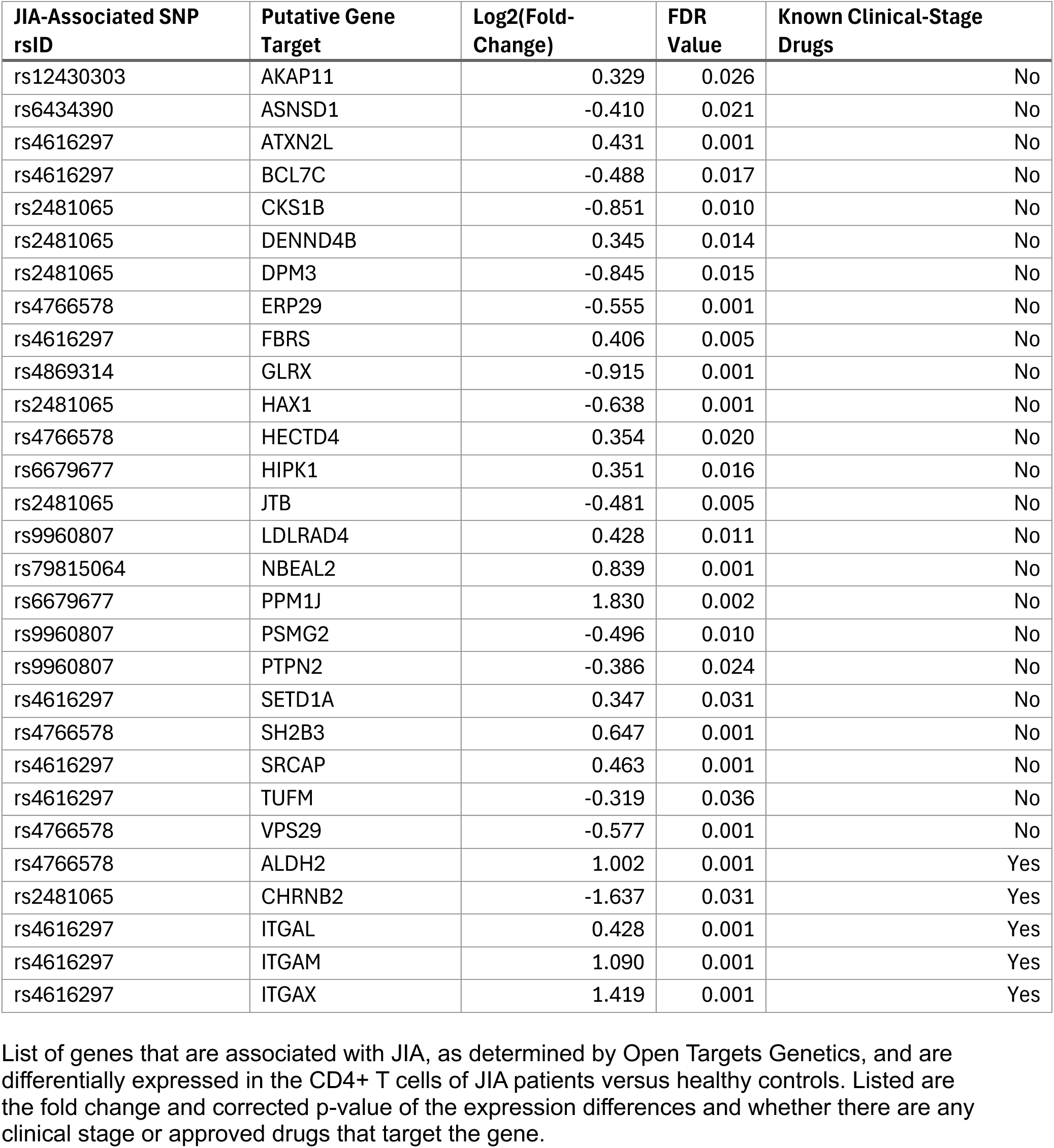
JIA Associated gene targets with differential expression.

**Table 2.** JIA associated genes with strong drug tractability.

| JIA-Associated SNP rsID | Putative Gene Target | Log2(Fold-Change) | FDR Value | Subcellular Localization |
| --- | --- | --- | --- | --- |
| rs12430303 | AKAP11 | 0.329 | 0.026 | Cytosol, Nucleoli, Plasma Membrane |
| rs2481065 | CKS1B | -0.851 | 0.010 | Cytosol, Mitochondria |
| rs4869314 | GLRX | -0.915 | 0.001 | Cytosol, Plasma membrane |
| rs6679677 | HIPK1 | 0.295 | 0.016 | Cytosol, nucleoplasm |
| rs9960807 | PTPN2 | -0.386 | 0.024 | Nucleoplasm |
| rs4766578 | VPS29 | -0.577 | 0.001 | Cytosol, vesicles |

List of genes that are associated with JIA, as determined by Open Targets Genetics, are differentially expressed in the CD4+ T cells of JIA patients versus healthy controls and had strong drug tractability for either small molecules or antibodies. Listed are the fold change and corrected p-value of the expression differences and their subcellular location.

**Table 3.** Predicted Gene Targets of rs6679677 JIA associated SNP.

| Gene | Overall L2G score | Partial L2G Scores |  |  |  |  |
| --- | --- | --- | --- | --- | --- | --- |
|  |  | Variant Pathogenicity | Distance | QTL Coloc | Chromatin Interaction | Distance to Locus |
| PTPN22 | 0.352 | 0.718 | 0.198 | 0.287 | 0.219 | 110567 |
| PHTF1 | 0.236 | 0.049 | 0.266 | 0.032 | 0.034 | 1697 |
| RSBN1 | 0.092 | 0.049 | 0.194 | 0.035 | 0.038 | 51290 |
| DCLRE1B | 0.035 | 0.064 | 0.030 | 0.277 | 0.155 | 143433 |
| HIPK1 | 0.030 | 0.064 | 0.029 | 0.243 | 0.155 | 168138 |
| BCL2L15 | 0.027 | 0.050 | 0.035 | 0.035 | 0.037 | 126395 |
| AP4B1 | 0.018 | 0.060 | 0.020 | 0.038 | 0.044 | 144015 |
| OLFML3 | 0.017 | 0.061 | 0.018 | 0.130 | 0.142 | 218205 |
| MAGI3 | 0.012 | 0.049 | 0.013 | 0.016 | 0.028 | 370671 |
| SYT6 | 0.010 | 0.049 | 0.009 | 0.032 | 0.034 | 392694 |

Putative target genes of the rs6679677 SNP, as defined by Open Target Genetics. Listed is the overall lead to gene score from Open Targets for each gene associated with the SNP, along with the partial L2G scores from the Open Targets model.

Differential expression analysis between the patients and healthy subjects revealed three genes that both met the expression criteria and were physically linked to the rs6679677 SNP, only one of which, *HIPK1*, was also linked to the SNP via OTG. The normalized gene expression values between disease states for *HIPK1*, showing lower expression in active disease compared to healthy controls, are shown in **Figure 2**. Overall, these data suggest that suppression of *HIPK1* in CD4+ T cells may be mediated by the rs6679677 SNP, which alters the regulatory activity of distal elements that influence *HIPK1* expression via physical proximity.

**Figure 2.**
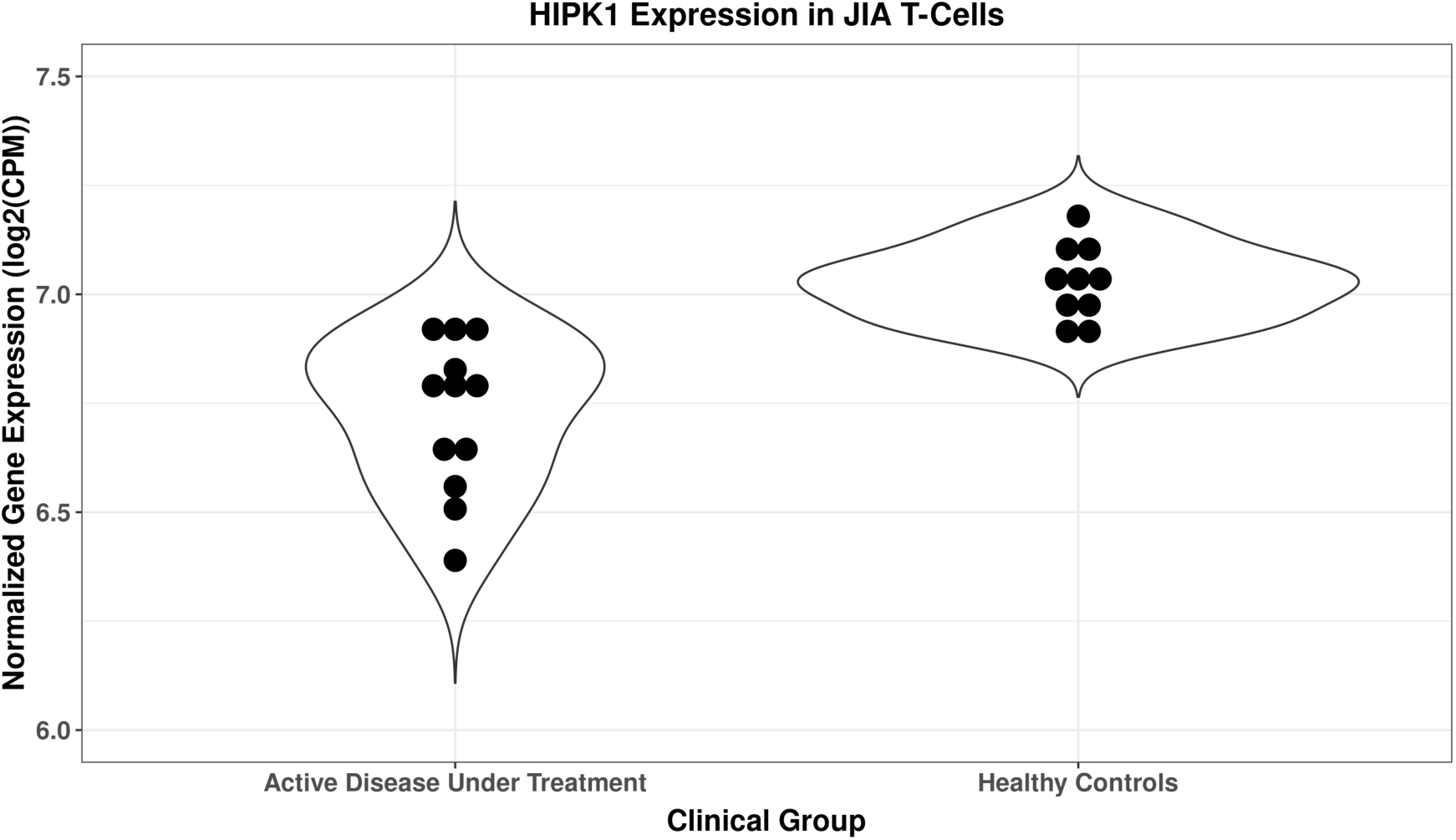
HIPK1 expression in T-cells of JIA patients and healthy controls, determined by RNA-seq, from Tarbell et al., 2021. Violin plots of HIPK1 normalized gene expression, measured as log2(counts per million), for healthy controls and patients with active disease and under treatment. Black dots represent individual level expression.

To elucidate the downstream events of *HIPK1* suppression in CD4 T cells and their role in the pathogenesis of JIA, additional analysis was performed. GO term analysis of the differentially expressed genes between active JIA and healthy controls revealed that of the top 15 most enriched terms, 4 of them were associated with *HIPK1*, indicating that the gene is involved in numerous processes that are altered in the disease state (**Table 4**).

**Table 4.** HIPK1 is involved in numerous critical processes.

| GO Term | Description | FDR Value |
| --- | --- | --- |
| **GO:0030155** | **regulation of cell adhesion** | **1.63E-03** |
| GO:0090287 | regulation of cellular response to growth factor stimulus | 8.76E-04 |
| GO:0006928 | movement of cell or subcellular component | 2.24E-03 |
| GO:0090092 | regulation of transmembrane receptor protein serine/threonine kinase signaling pathway | 5.73E-03 |
| GO:1903053 | regulation of extracellular matrix organization | 5.58E-03 |
| **GO:0050794** | **regulation of cellular process** | **5.93E-03** |
| GO:0090288 | negative regulation of cellular response to growth factor stimulus | 6.16E-03 |
| GO:0043087 | regulation of GTPase activity | 7.04E-03 |
| **GO:0023051** | **regulation of signaling** | **1.06E-02** |
| GO:0090101 | negative regulation of transmembrane receptor protein serine/threonine kinase signaling pathway | 1.03E-02 |
| GO:0016477 | cell migration | 1.26E-02 |
| **GO:0010646** | **regulation of cell communication** | **1.20E-02** |
| GO:0051056 | regulation of small GTPase mediated signal transduction | 1.17E-02 |
| GO:0007010 | cytoskeleton organization | 1.32E-02 |
| GO:1903391 | regulation of adherens junction organization | 1.26E-02 |

Top 15 GO terms enriched among the JIA down-regulated genes. Terms in bold represent terms that
are associated with HIPK1.

Understanding the role of different genes in the gene regulatory network of *HIPK1* may shed light on the role *HIPK1* is playing in JIA. To that end, the gene regulatory network for *HIPK1* was determined using String-DB, a repository of protein binding data (**Figure 3**). PML nuclear body scaffold protein, a binding partner of and coregulator with HIPK1, was found to be differentially expressed between active disease and healthy controls, in the same direction, but to a lesser degree, as *HIPK1* (**Figure 4**).

**Figure 3.**
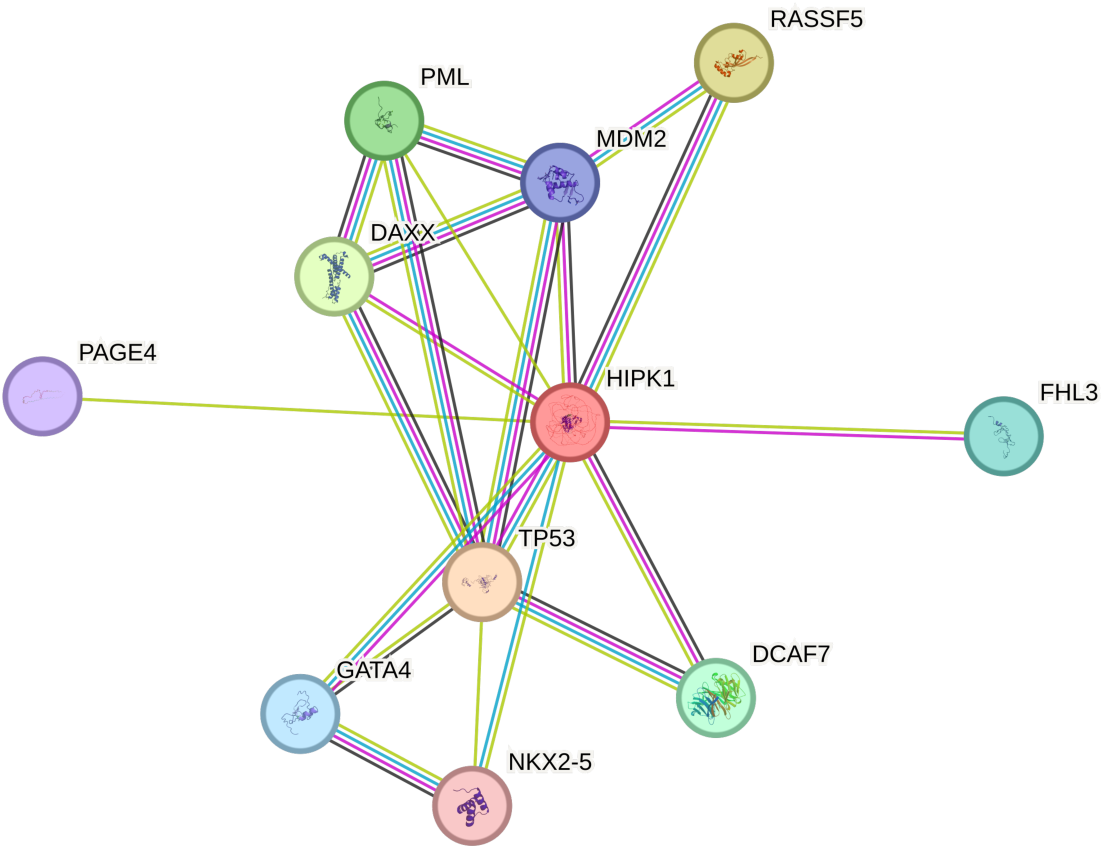
Gene Regulatory Network of HIPK1. Protein binding partners of HIPK1 as determined by StringDB. Green lines represent associations gleaned by textmining, purple lines represent results of direct binding experiments, blue lines represent associations curated from other databases and brown lines represent associations determined through coexpression.

**Figure 4.**
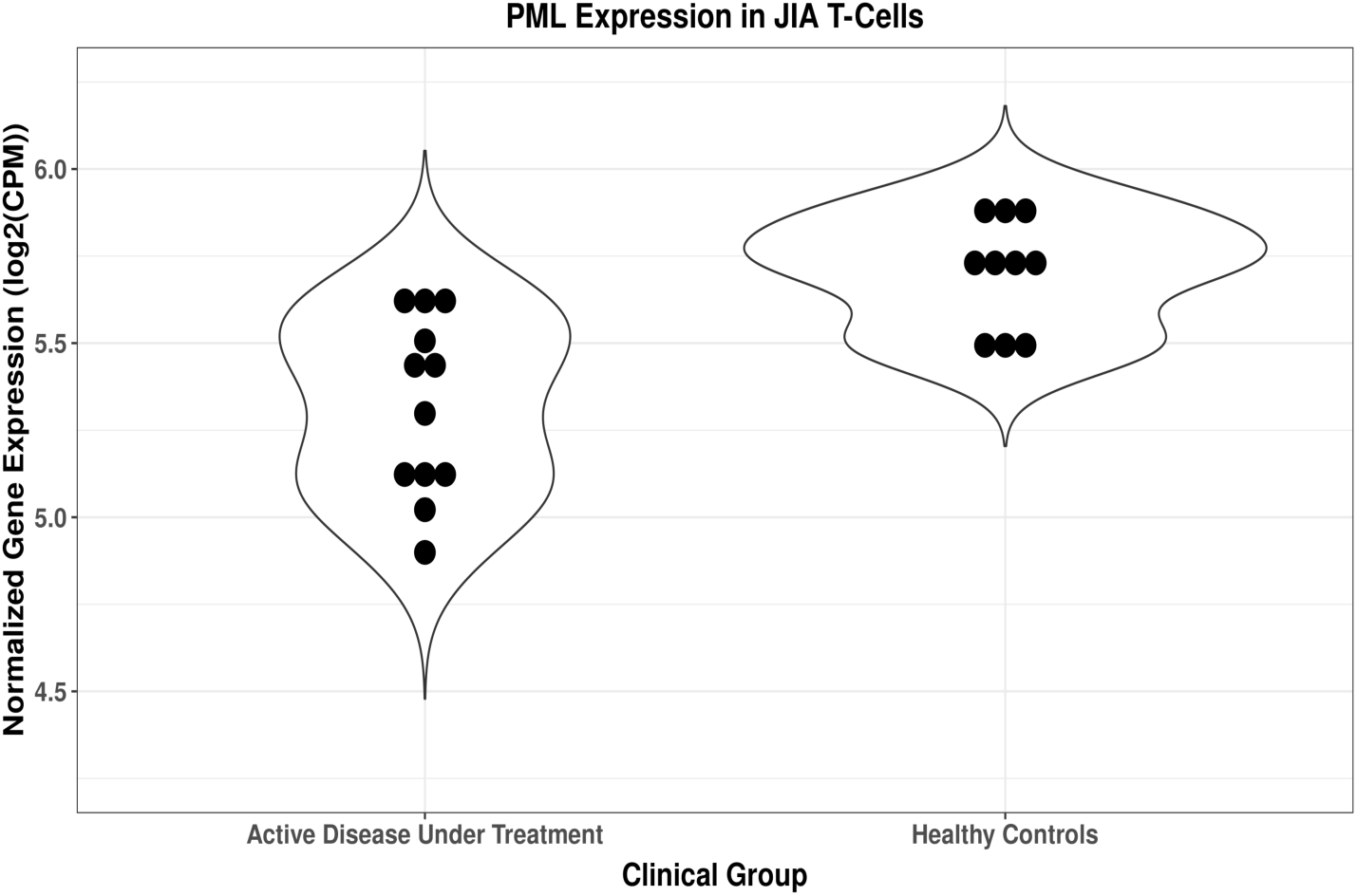
PML expression in T-cells of JIA patients and healthy controls, determined by RNA-seq, from Tarbell et al., 2021. Violin plots of PML normalized gene expression, measured as log2(counts per million), for healthy controls and patients with active disease and under treatment. Black dots represent individual level expression.

The remaining binding partners of HIPK1 in the gene regulatory network were crossed referenced to the FactorBook transcription factor repository, to determine which of the proteins acted as downstream transcription factors for HIPK1 signaling. Two factors were identified, p53 and GATA4. Transcription factor binding data, determined with ChIP-seq, were downloaded from FactorBook for both factors and run through BETA, using the differential gene expression dataset. As is shown in **Figures 5-7**, both factors show a strong statistical enrichment of binding sites nearby up-regulated genes, as opposed to down-regulated or non-differentially expressed genes.

**Figure 5.**
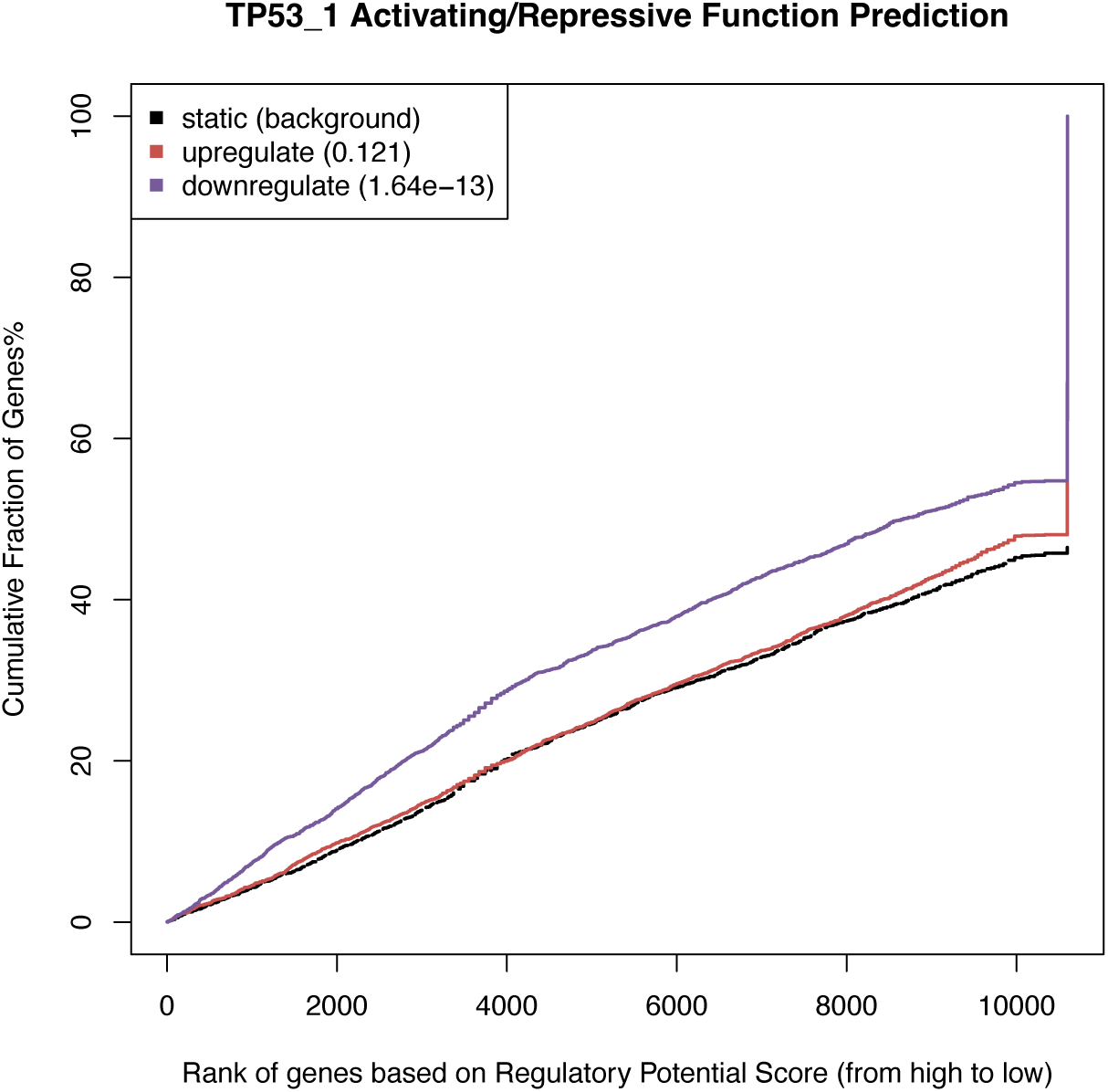
Regulatory Potential of TP53 ChIP-seq binding sites identified in A549 cells, for Differentially Expressed Genes in T-cells of JIA Patients. BETA generated cumulative distribution function of the gene groups and results of a one-tailed Kolmogorov-Smirnov test to determine whether the Up-regulated and Down-regulated groups differ significantly from the Non-differential group. The dotted line represents the background, the genes that are not differentially expressed, whereas the red and the blue lines represent the genes upregulated and downregulated, respectively. The y axis represents the proportion of genes in a category that are ranked at or better than the x-axis value, which represents the rank on the basis of the regulatory potential score from high to low. The P value listed in the top left represents the significance of the UP or DOWN group relative to the NON group as determined by the Kolmogorov-Smirnov test.

**Figure 6.**
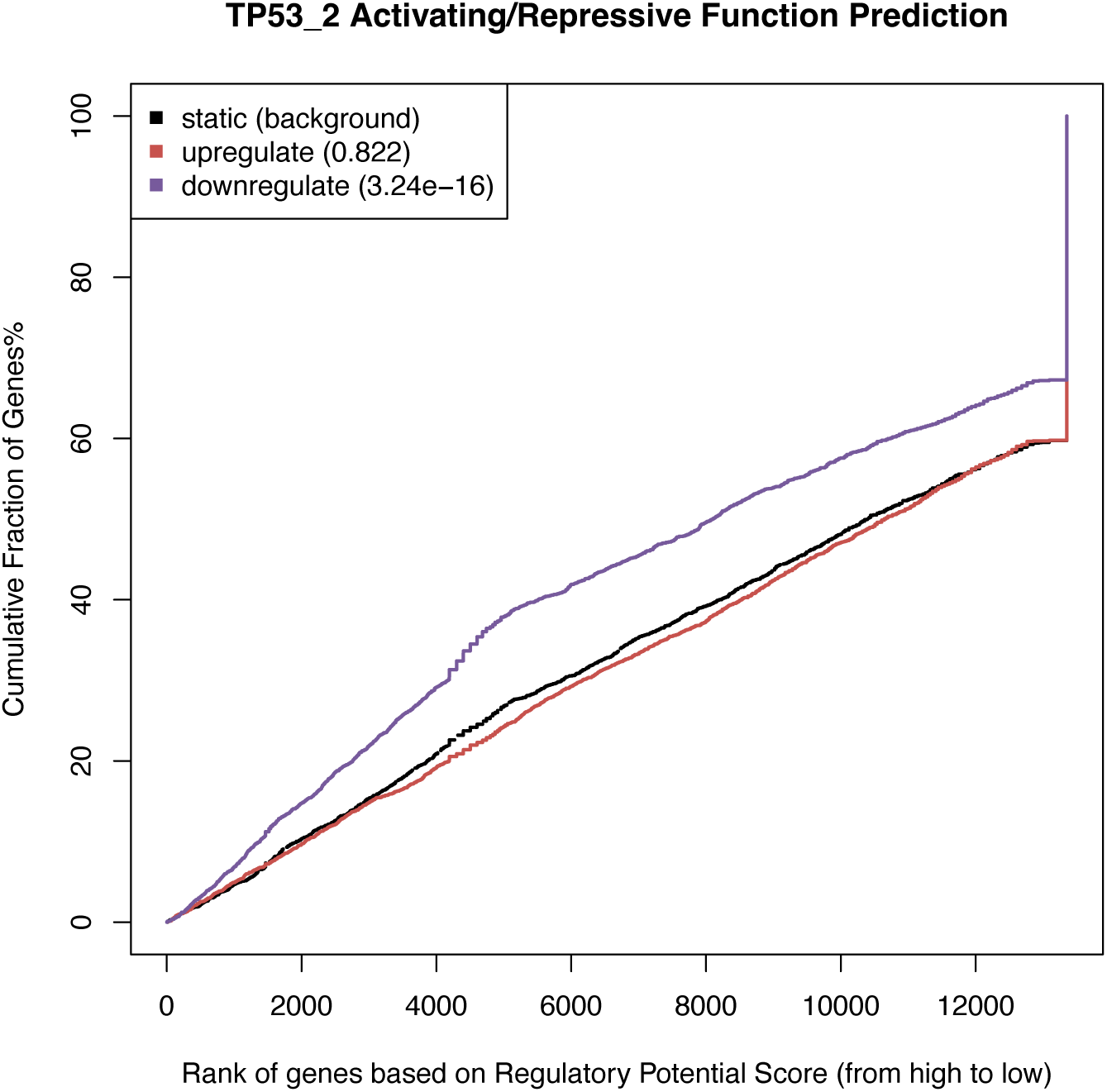
Regulatory Potential of TP53 ChIP-seq binding sites identified in HepG2 cells, for Differentially Expressed Genes in T-cells of JIA Patients. BETA generated cumulative distribution function of the gene groups and results of a one-tailed Kolmogorov-Smirnov test to determine whether the Up-regulated and Down-regulated groups differ significantly from the Non-differential group. The dotted line represents the background, the genes that are not differentially expressed, whereas the red and the blue lines represent the genes upregulated and downregulated, respectively. The y axis represents the proportion of genes in a category that are ranked at or better than the x-axis value, which represents the rank on the basis of the regulatory potential score from high to low. The P value listed in the top left represents the significance of the UP or DOWN group relative to the NON group as determined by the Kolmogorov-Smirnov test.

**Figure 7.**
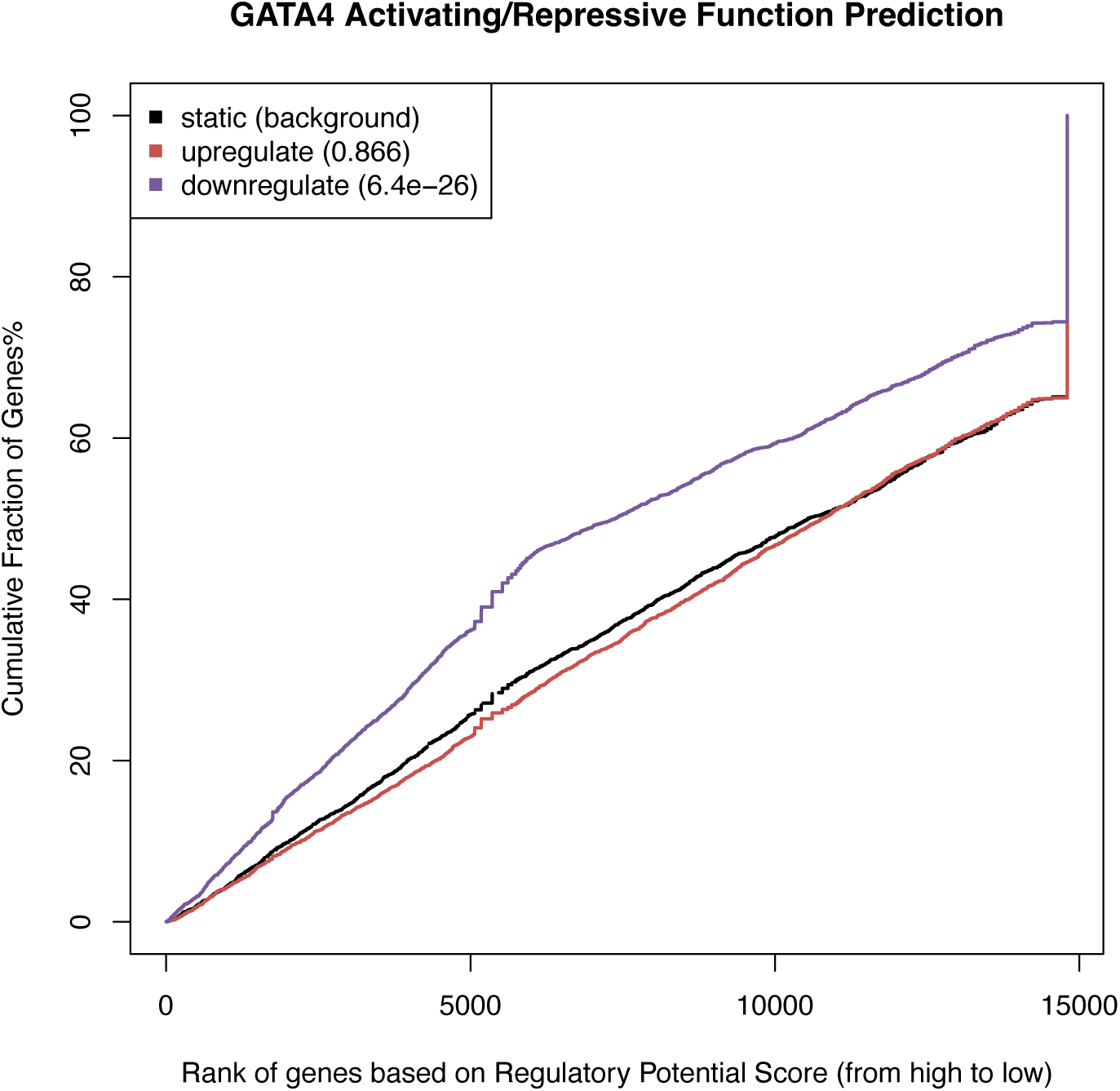
Regulatory Potential of GATA4 ChIP-seq binding sites identified in HepG2 cells, for Differentially Expressed Genes in T-cells of JIA Patients. BETA generated cumulative distribution function of the gene groups and results of a one-tailed Kolmogorov-Smirnov test to determine whether the Up-regulated and Down-regulated groups differ significantly from the Non-differential group. The dotted line represents the background, the genes that are not differentially expressed, whereas the red and the blue lines represent the genes upregulated and downregulated, respectively. The y axis represents the proportion of genes in a category that are ranked at or better than the x-axis value, which represents the rank on the basis of the regulatory potential score from high to low. The P value listed in the top left represents the significance of the UP or DOWN group relative to the NON group as determined by the Kolmogorov-Smirnov test.

## Discussion

Based on the findings presented here, we propose the following mechanism. In healthy CD4+T cells, under activated conditions, *HIPK1* plays a negative feedback role, acting to suppress GATA4 and TP53 and reduce their gene expression programs. In JIA, genetic variants in the vicinity of the rs6679677 index SNP result in the suppression of *HIPK1* expression, reducing the effectiveness of its regulatory role on GATA4 and TP53, prolonging a pro-inflammatory gene expression program that results in a hyper- and/or auto-inflammatory phenotype (**Figure 8**). This mechanism represents an ideal target for therapeutic intervention.

**Figure 8.**
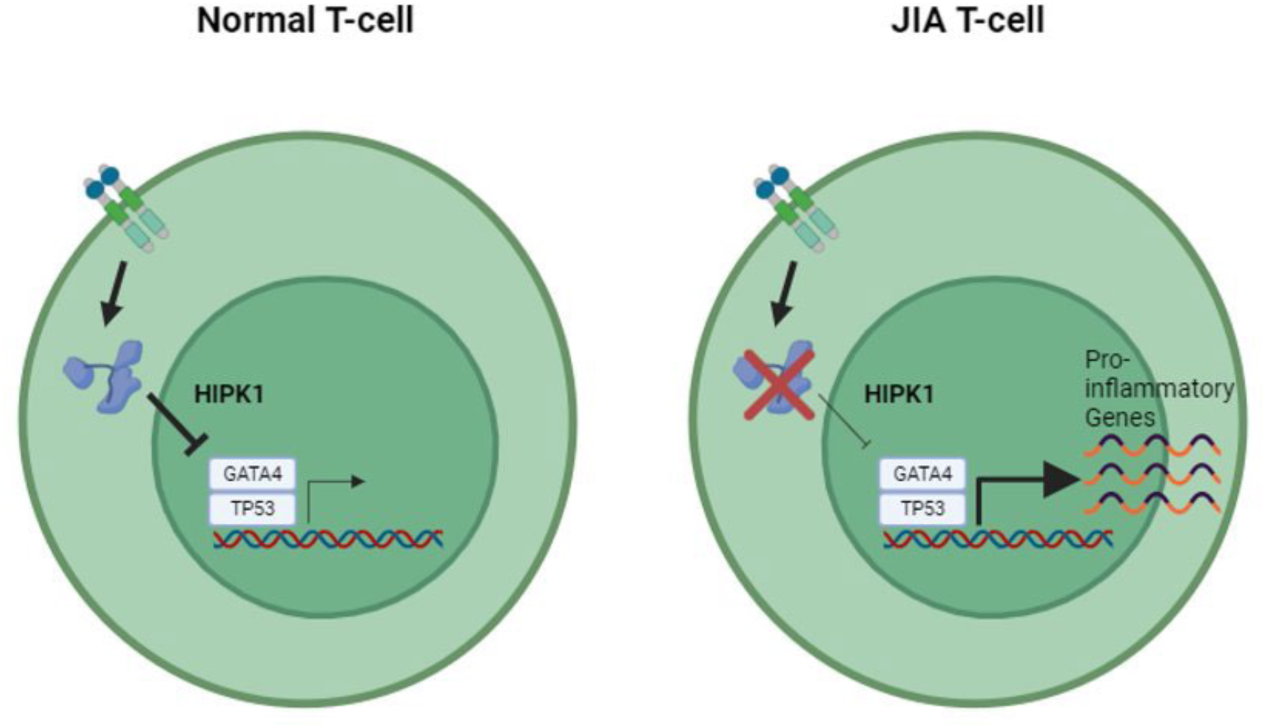
Model of HIPK1’s role in the pathogenesis of JIA. In healthy CD4+T cells, under activated conditions, HIPK1 acts as a negative feedback, acting to suppress GATA4 and TP53 binding and reduce their gene expression programs. In JIA, genetic variants in the vicinity of the rs6679677 index SNP reduce HIPK1 expression, lowering the effectiveness of its regulatory role on GATA4 and TP53 and prolonging a pro-inflammatory gene expression program that results in a hyper- and/or auto-inflammatory phenotype.

While the field of pediatric rheumatology has made significant advances in treating JIA in the past 20 years, significant challenges remain. For example, in a recent review using both Cincinnati Children’s Hospital Medical Center and Children’s Arthritis and Rheumatology Research Alliance Data (from a total of 8,978 patients), Brunner et al^10^ found that more than half the patients were treated with one or more biological agents, and that >10% of the patients continued to have active disease despite having been treated with ≥ 2 biological agents. Indeed, among those patients treated with ≥ 2 biological agents, more than half continued to experience active disease. Furthermore, the emergence of validated tools to capture patient-centered assessments of treatment outcomes^11^ is likely to reveal additional challenges not immediately obvious to physicians but unacceptable from a parent’s / patient’s point of view^3^.

A major impediment to developing improved, more precisely targeted therapies in JIA is our poor understanding of either the pathobiology of the disease or the biology of therapeutic response. With respect to the latter, we know that clinically derived descriptions of therapeutic response (active disease, inactive disease, clinical remission on medication) as established in the Wallace criteria^12^ have objective correspondents in peripheral blood gene expression profiles^13^. Therapeutic response is associated with complex re-organization of peripheral blood gene expression networks^14, 15^, but the most important elements of re-organization required to drive remission remain unknown. It is now recognized that remission is not typically associated with normalization of peripheral blood gene expression profiles^16^. However, while transcriptome studies have provided some useful information about the complex biology of treatment response, they have, by themselves, not led to identification of novel or unrecognized therapeutic targets.

In the current study, we used a combined genetics-genomics-transcriptomics approach to identify new therapeutic target candidates. The validity of our approach is demonstrated by the fact that we agnostically identified known therapeutic targets, e.g., cell-surface integrins CD11a and CD11b. Indeed, the anti-CD11a monoclonal antibody, Efalizumab, was used to treat autoimmune diseases, including psoriasis, psoriatic arthritis and was tested in rheumatoid arthritis (SuppTable1), before being withdrawn for safety issues. More recently, Schittenhelm et al. showed that altered expression patterns of CD11a and CD11b in dendritic cells was associated with inflammation regulation in adult rheumatoid arthrtis^32^.

In the current study, drug tractability was an important criterion for the selection process, and *HIPK1* is notable for being a member of a druggable family. However, as a Ser/Thr kinase family member, the modality being referenced is a kinase inhibitor, which, if targeted against HIPK1 in the context of JIA , would be predicted to exasperate the problem. As such, HIPK1 itself may not represent a suitable drug candidate for JIA, but rather an anchor point, around which a search for a more suitable upstream regulator or downstream effector can take place.

The findings presented here regarding HIPK1’s role in the pathogenesis of JIA have interesting therapeutic implications for other indications. Bei et al. recently showed that the inhibition of HIPK1 in cardiomyocytes was a novel therapeutic approach to treating hypertrophic cardiomyopathy (HTC)^33^. Based on the data presented here, however, such a treatment would run the risk of inducing similar phenotypic consequences to JIA. Moreover, given that the rs6679677 SNP is associated with other indications beyond JIA, such as type 1 diabetes^34^, rheumatoid arthritis^35^, and systemic lupus erythematosus^36^, it is possible that HIPK1 inhibition in the context of treating HTC may result in immune-modulated adverse reactions beyond synovial joints, in other tissues such as the pancreas. To avoid such adverse events, it may prove necessary to deploy the HIPK1 inhibitors using an active targeting approach, by attaching it to a monoclonal antibody (mAb) to form an antibody-drug conjugate (ADC) or to enclose it in a liposome and attach mAbs to the liposome surface to create an antibody conjugated liposome (ACL).

There are several limitations to this study. The first is that the wet lab and procedures here were performed exclusively with CD4+ T cells. Although these cells are known to be an important feature of the immunopathology of JIA^9^, there is compelling evidence that both neutrophils [ ^17 18^ ] and monocytes [ ^19 20^ ] are involved in both genetic risk and disease biology. Because there are small but significant differences in 3D chromatin architecture across cells and cell lines that may highlight additional target candidates. A second limitation is the fact that our method, like that of Fang et al [ ^21^ ], uses the GWAS-identified SNPs as the starting point for surveying genetic architecture. Because of the phenomenon of linkage disequilibrium, the SNPs that tag risk loci are not necessarily those that drive disease risk. It is probable that knowing the actual risk-driving variants and using higher-resolution maps of 3D chromatin (e.g., using MicroC^22, 23^) will allow the identification of additional targets and more robust precision medicine approaches to treating JIA.

In conclusion, we demonstrate the use of genetic, genomic, and transcriptomic data in CD4+ T cells to identify new therapeutic targets in JIA. Our approach, which identified genes within or downstream of the GATA4-p53 pathways as potential targets, also provides new insight into the pathobiology of JIA. The role of genetic variants in attenuating HIPK1-mediated inhibition of CD4+ T cell activation deserves further investigation.

## Data Availability

All data produced are present in the manuscript.

https://www.ncbi.nlm.nih.gov/geo/query/acc.cgi?acc=GSE164215

## Declarations

The authors declare no competing interests

---

**Supplemental Table 1.** Clinical Trials of putative gene targets.

| ClinicalTrials.gov<br>ID |
| --- |
| NCT00034203 |
| NCT00276250 |
| NCT00287118 |
| NCT00669214 |
| NCT00312026 |
| NCT00302445 |
| NCT00096980 |
| NCT00336973 |
| NCT00249808 |
| NCT00737763 |
| NCT00501709 |
| NCT00051662 |
| NCT00000278 |
| NCT02101008 |
| NCT03950830 |
| NCT00312819 |
| NCT00435435 |
| NCT02735577 |

Clinical trial identification numbers for drugs against one of the putative target genes for JIA.

